# Cost Impact of Chronic Care Management Services in a Large Multi-Specialty Practice: A Pragmatic Outcomes Study

**DOI:** 10.64898/2026.03.07.26347834

**Authors:** Bennett W. Clark, Jacob Webster, Souvik Chatterjee, Michael D. Finch

## Abstract

**Background:** Chronic Care Management (CCM) services represent an underutilized Medicare benefit with potential to reduce healthcare costs and improve care coordination for beneficiaries with multiple chronic conditions.

**Objective:** To evaluate the real-world impact of CCM services on healthcare expenditures and patient out-of-pocket costs in a large multi-specialty outpatient practice.

**Design:** Pragmatic retrospective cohort study comparing Medicare beneficiaries enrolled in CCM services versus eligible non-enrolled beneficiaries.

**Setting:** Large multi-specialty outpatient clinic in Alabama with 77 physicians across more than 20 specialties.

**Participants:** Treatment group (n=6,093) consisted of patients continuously enrolled in CCM services between January 1, 2024 and December 31, 2024. Comparison group (n=30,432) included eligible patients who were not enrolled in CCM services during the same period.

**Intervention:** Structured CCM program delivered by licensed practical nurses (LPNs) providing monthly telephone or video encounters focused on care plan implementation, medication reconciliation, care coordination, preventive health maintenance, and social determinants of health.

**Main Measures:** Per-member-per-year (PMPM) paid amounts and patient out-of-pocket expenditures, adjusted for age and sex differences.

**Results:** The CCM treatment group demonstrated 13.6% lower unadjusted healthcare costs compared to the comparison group ($96 vs. $110 PMPM). After adjusting for demographic differences, cost savings increased to 17.1% ($75 vs. $89 PMPM). Patient out-of-pocket expenses were 16% lower in the treatment group ($29 vs. $34 PMPM). These savings were achieved despite the treatment group being 2% older on average and including 10% more female beneficiaries—both factors typically associated with higher healthcare costs.

**Conclusions:** Implementation of structured CCM services in a real-world multi-specialty practice setting is associated with reductions in both total healthcare expenditures and patient out-of-pocket costs. These findings support CCM as a practical, cost-effective approach to chronic disease management that benefits patients.

## Introduction

Over the past decade, the Medicare program has increasingly recognized the importance of non–face-to-face care management services for patients with chronic conditions. In 2015, the Centers for Medicare & Medicaid Services (CMS) introduced reimbursement for outpatient longitudinal care management—known as Chronic Care Management (CCM)—for patients with multiple chronic conditions. This initiative represented a significant shift in payment policy, acknowledging the value of non–visit-based care coordination [1].

Despite this policy change, uptake of CCM services has remained limited. Multiple studies report that CCM claims are filed for fewer than 5% of Medicare beneficiaries, even though an estimated 85% of beneficiaries meet eligibility criteria [2,3]. For those who do receive CCM services, the average duration of support is modest, at approximately 4.3 months per year.

Evidence suggests that primary care clinicians deliver more than two-thirds of reimbursable care-coordination services [4]. This pattern aligns with a growing body of research indicating that non–face-to-face care management (NFFCCM)—which includes both CCM (longitudinal chronic-condition management) and Transitional Care Management (TCM, post-discharge care)—may shift healthcare utilization away from inpatient and emergency department (ED) settings toward outpatient care.

This shift has important implications for both cost and outcomes in the Medicare program. For example, in a cohort of beneficiaries with type 2 diabetes, receipt of NFFCCM was associated with fewer hospital admissions and ED visits, alongside increased outpatient encounters [5].

Similarly, analyses of TCM services demonstrate reductions in cost and mortality when delivered within 30 days of discharge [6]. CMS-commissioned evaluations of CCM further indicate that beneficiaries receiving CCM experience lower overall healthcare spending, with monthly per-beneficiary savings increasing from approximately $28 after 12 months of CCM to $74 after 18 months [7].

Nevertheless, important gaps remain in the evidence base. First, while some data suggest CCM can shift care to lower-acuity settings [5], limited evidence exists regarding its impact on total cost of care in broad primary-care populations, as opposed to disease-specific cohorts. Second, most prior research has grouped CCM and TCM together under the umbrella of NFFCCM, despite substantial differences in clinical scope. The specific contribution of CCM therefore remains under-studied. Third, the most comprehensive analyses of CCM’s impact are now nearly a decade old [7], underscoring the need for updated evidence as utilization patterns and reimbursement structures have evolved.

The present study aims to advance the literature in two ways. First, we examine the impact of CCM services alone—distinct from TCM—on healthcare utilization and expenditures in a large private primary-care practice setting. Second, we update and extend prior analyses by focusing on a general primary-care population of Medicare beneficiaries, rather than a single-disease cohort, within a real-world practice environment. By doing so, we assess how CCM functions in contemporary primary care and its potential to influence utilization across a broad set of patients with chronic conditions.

## Methods

### Study Design

The 2017 Centers for Medicare & Medicaid Services (CMS)–sponsored Mathematica evaluation of the Chronic Care Management (CCM) program primarily assessed program efficacy at the system level. Our study sought to extend this work by emphasizing the effectiveness of CCM in a real-world practice environment. Specifically, we employed a practical study design to examine differences in reimbursement patterns and patient out-of-pocket expenditures within a single CCM provider and a large multi-specialty outpatient clinic. Guided by the Pragmatic–Explanatory Continuum Indicator Summary (PRECIS) domains, we intentionally prioritized external validity and practice relevance over system-level cost analyses [12]. While the Mathematica evaluation has already demonstrated aggregate cost savings associated with CCM, our focus was directed toward the operational challenges and financial implications encountered in contracting with a CCM provider in routine clinical care.

### Study Setting

This study was conducted at a large, multi-specialty outpatient clinic in Alabama comprising 77 physicians across more than 20 medical specialties. The clinic provides ambulatory care to an adult population and maintains an integrated electronic health record (EHR) system that supports both clinical and administrative data collection. The clinic serves a geographically diverse patient population across multiple counties in Alabama, with a payer mix representative of the broader Medicare population in the southeastern United States.

### Study Population

Clinic patients continuously enrolled in the CCM program between January 1, 2024 and December 31, 2024 served as the treatment group (n = 6,093). Clinic patients who met the CCM eligibility criteria but were not enrolled during the same period served as the comparison group (n = 30,432). All patients included in the analysis maintained continuous enrollment in either Medicare Advantage or traditional Medicare Part B throughout the study period to ensure complete claims capture.

### The Wellbox CCM Program Patient Eligibility and Enrollment

Patients were identified through retrospective review of claims data from the preceding 12 months. Inclusion criteria required individuals to have at least two chronic conditions, as defined by ICD-10 codes corresponding to the Healthcare Cost and Utilization Project (HCUP)/Chronic Condition Indicator Refined (CCIR) list [8]. Eligible patients were further restricted to those enrolled in Medicare Advantage or traditional Medicare Part B, as these payers reliably reimburse for CCM services.

Eligible patients were contacted using a structured outreach protocol consisting of up to four telephone calls and two short message service (SMS) text messages, with a minimum of three business days between contact attempts. During outreach, patients received standardized counseling regarding the purpose of CCM, the communication process between the CCM nurse and their primary or specialty providers, and the presence of standard Medicare copayments.

Patients who consented to participate were formally enrolled in the Wellbox CCM program. Verbal consent was documented in the patient’s medical record, and a follow-up letter confirming enrollment was sent to each participant.

### CCM Clinician Training

All CCM services were provided by licensed practical nurses (LPNs) employed by Wellbox Inc, a third-party provider of remote clinical services. Twenty-nine LPNs participated in the program. Each completed a structured two-week didactic training followed by a six-week supervised shadowing period with a graduated increase in clinical workload. Approximately 25% of onboarding time focused on clinical content and disease management principles, 40% on information technology and systems training (including both the Wellbox CCM platform and the clinic’s EHR), and 35% on documentation standards and quality assurance. Clinicians were required to demonstrate proficiency in all domains prior to independent patient management.

Ongoing continuing education was provided monthly, with focus areas determined by quality metrics, emerging clinical guidelines, and feedback from supervising registered nurses.

### Intervention

Each LPN managed a target panel of approximately 200 patients; part-time clinicians maintained a minimum panel of 150 patients. Panel assignments were made based on patient preference, language concordance, and clinician expertise.

During the initial CCM encounter, clinicians reviewed the patient’s EHR and conducted a structured telephone or video visit to establish a primary diagnosis and comprehensive nursing-focused care plan. The initial assessment included review of past clinical history, current medications, recent hospitalizations or emergency department visits, and identification of self-management priorities. Care plans were developed using condition-specific templates aligned with evidence-based clinical practice guidelines. For example, in patients with hypertension, plans incorporated recommendations from the American Heart Association, behavioral support for physical activity, medication adherence counseling, education on medication side effects, and dietary modifications consistent with the DASH (Dietary Approaches to Stop Hypertension) diet.

Subsequent CCM encounters were conducted on a monthly basis, with a target of 20 minutes of clinical contact time per month per patient. These encounters focused on implementing and revising the individualized care plan. Structured interview modules guided assessment and intervention in five domains: (1) preventive health maintenance (including cancer screening, immunizations, and annual wellness visits); (2) coordination of care among multiple specialists and providers; (3) social determinants of health (including food security, housing stability, and transportation); (4) connection to community resources; and (5) medication reconciliation. Clinicians emphasized specific modules during encounters based on patient preference or clinical need.

CCM nurses maintained regular communication with patients’ primary care physicians and specialists through the shared EHR system. CCM nurses were trained to report significant changes in patient status, new symptoms, or medication concerns to the treating physician within 24 hours. Care plans were co-signed by the patient’s primary care physician and updated at least every 90 days or following any significant change in clinical status.

### Disenrollment Criteria

Patients were contacted up to three times per month by phone and three times via SMS for ongoing engagement. Participants who did not respond for three consecutive months were administratively disenrolled. Patients could also request voluntary withdrawal from the program at any time. Patients who were hospitalized or experienced significant health status changes remained enrolled and received enhanced outreach during recovery periods.

### Data Source and Analysis

Adjudicated final-action claims data for services delivered in 2024 were obtained from the multi-specialty outpatient group in which the Medicare beneficiaries were enrolled. Data were available for both the treatment and comparison groups and included information on actual reimbursement as well as patient out-of-pocket expenditures. Claims data were extracted from the clinic’s practice management system and included all billed services with dates of service between January 1, 2024 and December 31, 2024.

Analyses consisted of descriptive statistics and regression-adjusted estimates of group differences in reimbursement and patient out-of-pocket costs. Regression models included age and sex as covariates to adjust for demographic differences between groups. Given the large sample sizes and the complete enumeration of the study population rather than a probability sample, statistical significance testing (p-values and confidence intervals) was not reported. The data represent the complete set of claims for the study population and therefore do not constitute a sample, nor do they represent a draw from an infinite or superpopulation. As such, the assumptions underlying p-value interpretation are not met [9]. Instead, we report effect sizes and percentage differences as measures of the magnitude of association.

### Ethical Considerations

This study was conducted as a quality improvement initiative using de-identified administrative claims data. No patient identifiers were included in the analytic dataset. The study was determined to be exempt from institutional review board oversight under federal regulations for quality improvement activities.

## Results

### Participant Characteristics

Table 1 presents the sample size, age distribution, and sex composition of the Chronic Care Management (CCM) treatment and comparison groups. The treatment group included 6,093 unique patients, while the comparison group included 30,432 eligible but non-enrolled patients. On average, the treatment group was 2% older than the comparison group (mean age 74.5 years vs. 72.8 years) and included 10% more female beneficiaries (62% vs. 56%). These demographic differences are noteworthy given that both advanced age and female sex are established risk factors for higher healthcare utilization and costs in the Medicare population [10,11].

**Table 1.** Characteristics and Outcomes of the Comparison and Treatment Groups.

| Characteristic | Comparison Group | Treatment Group | % Difference |
| --- | --- | --- | --- |
| Number of Unique Patients | 30,432 | 6,093 | — |
| Mean Age (years) | 72.8 | 74.5 | +2.3% |
| Female (%) | 56% | 62% | +10.7% |
| PMPM Paid Amount (unadjusted) | \$110 | \$96 | −13.6% |
| PMPM Paid Amount (adjusted for age and sex) | \$89 | \$75 | −17.1% |
| PMPM Patient Responsibility | \$34 | \$29 | −16.0% |

*Note: PMPM = per member per year. Adjusted estimates derived from linear regression models controlling for age and sex.*

### Cost Outcomes

With respect to reimbursement, measured as paid amounts per member per year (PMPM), the treatment group was 13.6% less expensive than the comparison group ($96 vs. $110 PMPM). Patient out-of-pocket expenditures were likewise 16% lower in the treatment group ($29 vs. $34 PMPM).

After adjusting for age and sex differences through regression modeling, the cost advantage of the CCM treatment group increased to 17.1% ($75 vs. $89 PMPM). This adjustment accounts for the fact that the treatment group’s demographic profile would typically predict higher, not lower, healthcare costs. The adjusted estimate therefore represents the isolated effect of CCM enrollment on healthcare expenditures, independent of baseline demographic risk.

Table 2 summarizes differences in care by place of service. The traditional drivers of health care costs for this population include Inpatient Hospital, On Campus–Outpatient Hospital, Emergency Room – Hospital, and Assisted Living Facilities.

**Table 2:** Percent of claims and Reimbursement by Place of Service.

| Table 2: Percent of claims and Reimbursement by Place of Service |  |  |  |  |  |  |  |  |  |
| --- | --- | --- | --- | --- | --- | --- | --- | --- | --- |
|  | Comparison Group |  |  | Treatment Group |  |  | percent Difference<sup>1</sup> |  |  |
| Place of Service | Percent of Claims | % of Reimbursement | Average Reimbursement | Percent of Claims | % of Reimbursement | Average Reimbursement | Percent of Claims | % of Reimbursement | Avg Reimbursement |
| Inpatient Hospital | 0% | 0% | \$ 36 | 0% | 0% | \$ 25 | | | -48% |
| Other Place of Service | 1% | 2% | 351 | 0% | 1% | \$ 302 | | | -16% |
| Emergency Room – Hospital | 4% | 6% | \$ 158 | 2% | 3% | \$ 139 | | | -13% |
| Office | 78% | 70% | \$ 98 | 88% | 83% | \$ 91 | 11% | 15% | -7% |
| On Campus-Outpatient Hospital | 14% | 18% | \$ 139 | 9% | 13% | \$ 130 | -49% | -41% | -7% |
| Ambulatory Surgical Center | 0% | 0% | \$ 123 | 0% | 0% | \$ 135 | | | 9% |
| Telehealth Provided Other than in Patient's Home | 0% | 0% | \$ 39 | 0% | 0% | \$ 51 | | | 25% |
| Telehealth Provided in Patient's Home | 0% | 0% | \$ 59 | 0% | 0% | \$ 80 | | | 25% |
| Assisted Living Facility | 0% | 0% | \$ 155 | 0% | 0% | \$ 210 | | | 26% |
| Missing | 3% | 4% | \$ 144 | 0% | 1% | \$ 246 | | | 42% |

1 % difference not shown for differences of 2% of less

Compared to the Comparison Group, the Treatment Group had 11% more claims for Office services but 49% fewer claims for On Campus–Outpatient Hospital. Office visits represented 15% of the Treatment Group’s total reimbursement and were 7% less expensive than the Comparison Group’s office visits. Similarly, use of On Campus–Outpatient Hospital services was 49% lower in the Treatment Group, accounted for 41% less of their reimbursement, and was on average 7% less expensive.

There are other instances where the Comparison Group outperformed the Treatment Group; however, the number of claims in these categories is extremely small—often less than one percent in both groups.

### Supplementary Findings

Among the treatment group, the average duration of CCM enrollment was 11.2 months (range: 12–12 months for continuously enrolled patients), with a mean of 9.8 clinical encounters per patient over the study period. Voluntary disenrollment occurred in 4.2% of enrolled patients, while administrative disenrollment due to non-engagement occurred in 3.1% of patients. The remaining 92.7% of enrolled patients maintained continuous engagement throughout the study period.

## Discussion

This retrospective cohort study demonstrates a substantial reduction in total healthcare expenditure and patient out-of-pocket costs for Medicare beneficiaries participating in CCM. These cost savings are more pronounced when controlling for age and gender. Our findings are consistent with an earlier generation of research on the impact of CCM on healthcare spending, in particular the findings of Schurrer et al. [7]

Our study provides several important updates to the existing literature on the impact of CCM on healthcare spending. First, it extends those findings to healthcare environments that more closely reflect contemporary healthcare delivery. In Schurrer’s foundational analysis of the impact of CCM on healthcare spending, the majority of CCM claims were filed by practices with five or fewer providers. Since that analysis was published, the American healthcare delivery system has undergone consolidation, with primary care increasingly provided by organizations with health systems and corporate owners. [13] Our results suggest that provision of CCM is associated with reduction in Medicare spending when delivered within a large, hospital-affiliated health system, as opposed to a large number of small, unaffiliated practices.

Our study extends prior research by demonstrating that favorable financial outcomes are associated with CCM services provided by clinicians not directly employed by the billing provider. The LPNs providing CCM services in this study had access to health system EMR and other health records and were able to communicate with primary care providers and other clinicians involved in their patient’s care. However, the CCM LPNs were not employed by the health system billing for their services and were not geographically collocated with their patient population. The Wellbox CCM program’s ability to shift healthcare access to lower-acuity sites and to reduce overall spending despite this disparity between primary provider and CCM clinician may be due to several factors. These include the dedicated CCM training LPNs received at the time of onboarding, customized clinical templates designed specifically for CCM, and robust training and integration between the vendor’s and healthcare system’s information management systems.

### CCM Impact on Site of Care

In our study, Medicare beneficiaries receiving CCM had increased rates of office-based professional healthcare services and lower utilization of hospital-based services—both inpatient and outpatient. While the use of inpatient services was small in both groups, the 48% lower reimbursement for the Treatment Group suggests a lower acuity level at the time of admission for patients in that group. This shift toward increased outpatient utilization likely contributed to the CCM cohort’s decrease in healthcare spending. Numerous prior studies have documented a link between increased outpatient primary care access and decreased total healthcare spending. [14–16] This body of research points toward the ability of outpatient care, and especially primary care, to prevent the decompensation of chronic disease and effectively coordinate care for patients with complex conditions. Against this background, the CCM services provided in this study can be seen as contributing to reduced cost of care in at least two ways. First, CCM extends core primary care functions associated with improved disease management, such as medication reconciliation and self-management of chronic disease. Second, CCM promotes patient engagement with outpatient and primary care providers [5], enabling the prevention of acute decompensation of chronic conditions.

### Policy Implications

Low national uptake of CCM has been well documented [2,3], with outpatient providers citing administrative complexity, documentation requirements, and staffing constraints as key barriers. Our study offers a pragmatic solution by evaluating a delivery model in which provider groups contract with a specialized third-party vendor to operationalize CCM. This model substantially reduces the logistical burden on outpatient practices: the vendor assumes responsibility for staffing, training, workflow management, and compliant documentation, allowing clinicians to offer CCM without diverting scarce internal resources. In turn, providers retain the ability to bill for CCM services—creating a new revenue stream that aligns financial and clinical objectives. By demonstrating meaningful reductions in total healthcare spending within this structure, our findings highlight an implementation strategy that could remove some of the most persistent barriers to CCM adoption.

Prior research on CCM has largely focused on disease-specific cohorts, particularly diabetes, heart failure, and other chronic conditions with high morbidity. Our study extends this work by demonstrating that CCM can generate broad cost benefits at the population level, rather than only within narrowly defined disease groups. This suggests that healthcare systems need not silo CCM programs by clinical specialty or reserve them exclusively for high-cost cohorts. Because our study’s CCM intervention was implemented at scale under real-world conditions, the results carry strong relevance for contemporary health systems, many of which face increasing pressure to deploy care-management interventions rapidly and consistently across large, distributed networks.

### Study Limitations and Future Research

This study analyzes only a single year of Medicare claims following CCM implementation. While this window is sufficient to detect early utilization and spending effects, it does not capture potential longer-term trajectories, such as whether savings persist, whether benefit wanes over time, or whether delayed adverse consequences emerge. Similarly, the reliance on claims data limits our analysis to spending outcomes; we did not evaluate clinical quality measures such as adherence, disease control, hospital readmission rates, or ambulatory care–sensitive conditions. Without such measures, the mechanisms by which CCM reduces spending remain incompletely understood, and we cannot exclude the possibility that reduced spending may in some cases reflect under-utilization rather than more efficient care.

Our study does not assess patient experience or patient-reported outcomes, an important omission given that CCM requires monthly copays for many Medicare beneficiaries. Copays, varying levels of engagement, and differences in communication preferences may meaningfully influence CCM utilization and effectiveness.

Future investigations should employ prospective or quasi-experimental designs to more rigorously evaluate the causal relationship between CCM exposure and cost reduction. Randomized or stepped-wedge designs would help disentangle CCM’s true effect from unobserved confounding and selection bias. Research should also examine more granular utilization patterns—such as specific sites of care, patterns of outpatient engagement, diagnostic categories, and subtypes of avoidable hospitalizations—to clarify where CCM achieves the greatest impact and for which patients.

Additional work should evaluate CCM’s patient experience, including perceived value, barriers to engagement, and the effect of copays on uptake. Such insights are essential for informing reimbursement policy and optimizing equity in program outreach. Finally, future research should assess program fidelity, workforce models, and differential effects across health-system contexts. Understanding how CCM performs across urban vs rural settings, integrated delivery systems vs independent practices, and medically complex vs relatively healthy populations will help healthcare leaders and policymakers target resources toward the environments where CCM is most likely to succeed.

## Conclusion

Chronic Care Management was associated with meaningful reductions in total healthcare spending, even after adjusting for demographic differences that would ordinarily predict higher costs. These findings reinforce the growing evidence that structured, non–face-to-face care management can shift utilization away from higher-acuity settings and toward lower-cost outpatient care.

The study’s pragmatic design—implemented within a large, contemporary health system and delivered through a contracted third-party vendor—offers a scalable, generalizable template for CCM adoption. By reducing operational burden while preserving clinical integration, this model provides a viable path for broader uptake and system-level cost containment.

## Acknowledgments

The authors wish to acknowledge the physicians, staff, and administrators of the participating multi-specialty clinic for their collaboration and support of this research. We also recognize the Wellbox Inc. care team for their dedication to delivering high-quality chronic care management services.

## Data Availability Statement

The data analyzed in this study contain protected health information and are not publicly available. De-identified aggregate data may be made available from the corresponding author upon reasonable request and with permission from the data-holding institution.

## Author Contributions

Conceptualization: BC

Data Curation, Formal Analysis: MF

Writing – Original Draft Preparation: BC, MF

Writing – Review & Editing: JW, SC, MF

